# Artificial intelligence-driven meta-analysis of brain gene expression identifies novel gene candidates and a role for mitochondria in Alzheimer’s Disease

**DOI:** 10.1101/2022.02.02.22270347

**Authors:** Caitlin A. Finney, Fabien Delerue, Wendy A. Gold, David A. Brown, Artur Shvetcov

**Author notes:** Authors to whom correspondence should be addressed. Caitlin A Finney: 176 Hawkesbury Rd. Westmead, NSW, Australia, and Artur Shvetcov: Hospital Rd. Randwick, NSW, Australia. These authors contributed equally to the work (Caitlin A. Finney, Artur Shvetcov).

## Abstract

Alzheimer’s disease (AD) is the most common form of dementia. There is no treatment and AD models have focused on a small subset of genes identified in familial AD. Microarray studies have identified thousands of dysregulated genes in the brains of patients with AD yet identifying the best gene candidates to both model and treat AD remains a challenge. We performed a meta-analysis of microarray data from the frontal cortex (n = 697) and cerebellum (n = 230) of AD patients. A two-stage artificial intelligence approach, with both unsupervised and supervised machine learning, combined with a functional network analysis was used to identify functionally connected and biologically relevant novel gene candidates in AD. We found that in the frontal cortex, genes involved in mitochondrial energy, ATP, and oxidative phosphorylation, were the most significant dysregulated genes. In the cerebellum, dysregulated genes were involved in mitochondrial cellular biosynthesis (mitochondrial ribosomes). There was little overlap between dysregulated genes between the frontal cortex and cerebellum. A further functional network analysis of these genes identified that two downregulated genes, ATP5L and ATP5H, which both encode subunits of ATP synthase (mitochondrial complex V) may play a role in AD. Combined, our results suggest that mitochondrial dysfunction, particularly a deficit in energy homeostasis, may play an important role in AD.

## 1. Introduction

Alzheimer’s disease (AD) is the most common form of dementia with 50 million suffering from it globally and almost 10 million people developing it yearly ^1^. There are currently no effective treatments for AD and over 99.6% of clinical trials of AD have failed so far ^2^. Further, most of what is currently known about AD has been established using familial AD (FAD) models, characterized by mutations in amyloid precursor protein (APP), presenilin 1 (PSEN1) and presenilin 2 (PSEN2), which account for less than 1% of AD cases ^3, 4, 5, 6^.

The recent advent of high-throughput genetic analysis technologies have allowed us to delve into the dysregulated genetic landscape of sporadic, or late onset, AD. Gene expression profiling via microarray analysis allows quantitation of a large number of mRNA transcripts and their variation in a relatively unbiased approach ^7^, but have previously been confined to relatively small sample numbers, making statistical analysis challenging ^8^. More specifically, the dataset is pruned using arbitrary fold changes as well as p-values that require multiple corrections, both of which potentially overlook genes that may be important. Additionally, there is a tendency to use microarray data to confirm *a priori* hypotheses, perhaps biasing reported outcomes rather than using microarray as a purely exploratory method.

Recent meta-analyses of publicly available microarray data have proven to be a rich vein for identifying novel genes contributing to several diseases, including influenza ^9^, atherosclerosis ^10^, chronic pain ^11^, cancer ^12, 13^, and Parkinson’s disease ^14^. To date, however, few studies have used this approach to investigate possible genetic contribution to AD. One such study took a comparative approach to find overlapping gene expression profiles in neurodegenerative disorders including AD, Lewy body disease, amyotrophic lateral sclerosis and frontotemporal dementia ^15^. Another study took a similar approach to compare cross-species transcriptional overlap between mouse models of AD and humans ^16^. These meta-analyses, however, are limited in two ways. First, human transcriptomic data is high-dimensional (i.e. high ratio of samples to genes) and complex, making it difficult to identify disease associated patterns in the datasets ^17^. Second, as discussed above, these studies identified thousands of dysregulated genes without identifying which of these may be the best target(s) for developing new mouse and cell models of AD to interrogate and identify novel therapeutic approaches.

The use of artificial intelligence (AI) is a way to overcome the above-mentioned limitations. Importantly, not only is AI able to unravel patterns within complex data in an unbiased way ^17^, it also has the ability to reveal which gene target(s) should be investigated further. An example of this is that machine learning models have successfully differentiated Parkinson’s disease patients from healthy controls based on their gene expression profiles ^18^. Further, a recent study using a similar approach, in inflammatory bowel disease (IBD), performed a meta-analysis of human gene expression data, followed by machine learning, to identify novel disease causing genes ^19^. Their role in disease was confirmed in mice, representing the development of a novel model of IBD. Using these recently developed tools, patient-derived IBD organoids were successfully ‘treated’, identifying novel therapeutic targets and therapies for IBD ^19^. In AD, as far as we are aware, a meta-analysis combined with machine learning approach has been used once. However the identified genes had little to no functional interactions in STRING pathway analysis, suggesting that they are unlikely to be biologically relevant ^20^. In the context of AD, these previous data establish a clear need to for methods that identify functionally connected and biologically relevant candidate genes as has been done in IBD ^19^. Hopefully, this approach will lead to the development of novel animal models of AD as well as new treatments.

To identify the most biologically relevant genes in AD that inform disease pathophysiology, we performed a meta-analysis of an unprecedented number of microarray datasets from the frontal cortex and cerebellum of patients with AD compared to healthy controls. We then used a combination of unsupervised and supervised machine learning along with functional network analyses in STRING to determine genes with clearly established interaction networks indicating that that may be biologically relevant to AD.

## 2. Methods

### 2.1. Systematic Review of Publicly Available Data Repositories

To identify publicly available transcriptomic datasets, a systematic review of the Gene Expression Omnibus (GEO) database was performed. The key search terms used included “Alzheimer’s disease” and “homo sapiens”. Datasets were screened based on the following inclusion criteria: (a) gene expression data generated using microarray platforms, (b) gene expression specific to the amygdala, hippocampus, entorhinal cortex, frontal cortex, temporal cortex or cerebellum, (c) clinically confirmed Alzheimer’s disease patients and (d) inclusion of cognitively normal healthy controls. Datasets were excluded for the following: (a) use of other high throughput gene expression assays (e.g. RNA-sequencing), (b) gene expression in the periphery (e.g. blood), (c) not specifying confirmed AD diagnosis, and (d) not including cognitively healthy controls (e.g. use of controls with mild cognitive impairment).

### 2.2. Identification and Meta-Analysis of AD Microarray Datasets

A total number of 1010 datasets in GEO were screened. Based on the inclusion and exclusion criteria, 14 datasets were identified as being eligible for inclusion in the meta-analysis. The meta-data of each dataset was analyzed to determine if there was a sufficient sample size to undertake further analyses and machine learning. Of these datasets, 11 were excluded due to an insufficient sample size and were unable to be combined to create a new dataset of sufficient size (e.g. sample size of only 10). Consequently, we were unable to include the following brain regions in the analyses: the amygdala, hippocampus, entorhinal cortex, and temporal cortex.

In the three identified data sets, we included two brains regions, the frontal cortex and cerebellum. Data from the frontal cortex originated from two GEO datasets: GSE44770 and GSE33000. Data for the cerebellum originated from the GEO dataset GSE44768. Samples from the studies consisted of AD patients, with confirmed antemortem clinical diagnosis and postmortem neuropathological assessment, and normal, non-demented, healthy controls that were age-matched ^21, 22^. After identifying the respective datasets for the frontal cortex and cerebellum, we then converted all normalized gene expression values into a z-score. For the frontal cortex only, genes common to both datasets were selected and then the two datasets were merged into one. Data processing and merging was done in R Studio v1.2.5033 (R v3.6.3) with packages GEOquery, and dplyr. The final, unified frontal cortex dataset had a total sample size of n = 697 (AD n = 439, healthy control n = 238). The cerebellum dataset had a total sample size of n = 230 (AD n = 129, healthy control n = 101).

### 2.3. Machine Learning and STRING Network Analysis

The conventional approach for identifying differentially expressed genes typically uses fold change, *p*-values, or a combination of the two. These methods, however, are limited and are unlikely to provide the information needed to make strong conclusions about dysregulated genes that may be important for AD. First, using standard fold change cutoffs to select differentially expressed genes is inherently problematic. Genes with either low, or high, absolute expression are more likely to either easily meet or miss the fold change threshold, respectively, regardless of whether or not the gene is truly differentially expressed ^23, 24^. Further, the *p*-value is well-known to not only provide limited information about the data at hand, but to also be easily misinterpreted, thus likely contributing to the replication crisis ^25, 26, 27^. Specifically, calculating statistical significance using a *p*-value does not account for the degree to which the genes are, or are not, involved in differences between groups (in this case, between AD patients and healthy controls). One way to circumvent the inherent limitations of conventional methods is to approach the problem of identifying novel disease-driving genes through the lens of artificial intelligence, such as machine learning. In other words, we treat the problem as a machine learning problem: we would like to determine which genes best predict the classification of a given sample as being from either an AD patient or healthy control.

In the present study, we used a two-stage machine learning approach (Figure 1). In the first stage, we used unsupervised machine learning to perform an initial feature selection: identifying the genes that are likely to be important candidates for distinguishing an AD patient from a healthy control. Feature selection is an important step to reduce the number of features and thus avoid the curse of dimensionality for the final machine learning algorithm ^28^. Unsupervised machine learning is unique in the sense that it does not predefine any sample as being either an AD or healthy control sample. Instead it identifies similarities between the various samples that exist, irrespective of their group membership ^29^. Samples with high similarity will cluster together and will inform us how the data is grouped and what the drivers of this differentiation are ^29^. If true differences exist between AD patients and healthy controls, the unique clusters will be representative of this, and we will identify which genes are driving these differences. The unsupervised machine learning approach used here was a principal component analysis (PCA), an important technique that reduces dimensionality within a dataset while simultaneously minimizing any information loss ^30^. PCA has an established use in the analysis of high throughput datasets, such as microarray, to reveal hidden patterns within the thousands of identified genes ^31, 32, 33^. All the genes (≥ 15,000 genes per dataset) that were identified within the frontal cortex and cerebellum datasets, respectively, were analyzed using PCA in R Studio v1.2.5033 (R v3.6.3) and visualized using ggplot (Figure 1). The PCA indicated that the between group variance for the frontal cortex ran along principal component (PC) 1 and along PC2 for the cerebellum. As such, the top 1000 genes that contributed to frontal cortex PC1 and cerebellum PC2, respectively, were selected as potential gene candidates for AD (Figure 1).

**Figure 1.**
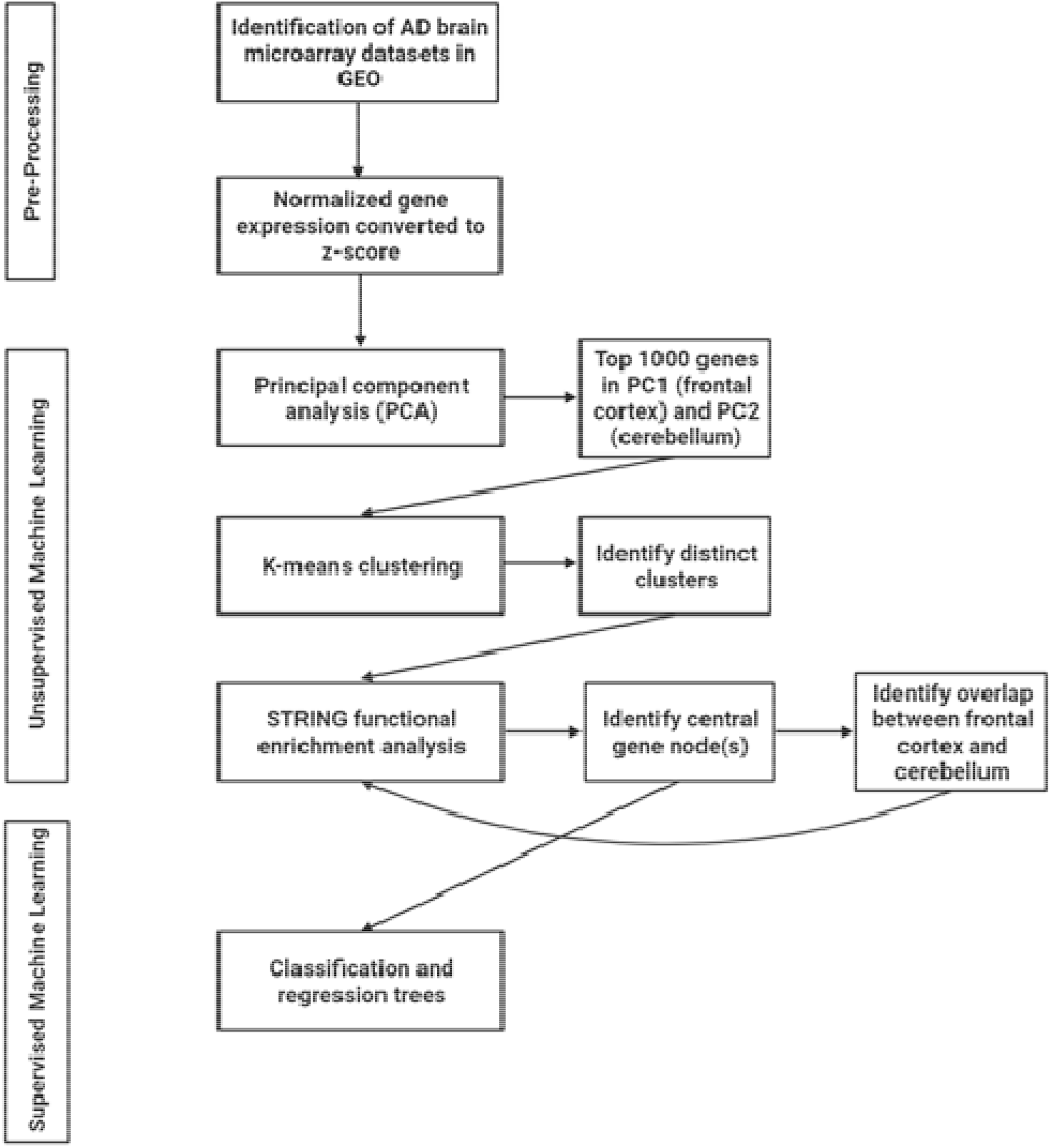
AI workflow used in the current study to identify new AD related genes.

To further narrow down the list of gene candidates, we entered each list of the top 1000 contributing genes from the PCs into STRING v11 ^34^. Importantly, STRING allows for the identification of interaction networks and gene-enrichment analysis ^34^. We then identified possible distinct network clusters using k-means clustering in STRING (Figure 1). K-means clustering is an unsupervised machine learning approach which, here, is a way to identify genes in the network that have similar interconnections and overlapping pathways ^35^. Subsequently, each distinct k-means cluster for the two brains regions were separately entered into STRING and a network analysis was performed using the following active interaction sources: experiments, databases, co-expression, neighborhood, and gene fusion and a minimum required interaction score of 0.7 (high confidence) (Figure 1). Importantly, using STRING, and the wealth of experimental data that it relies on, to identify pathways and interaction networks increases the likelihood of identifying strong, biologically relevant gene candidates for AD. More specifically, this approach allows for the identification of biologically relevant pathways rather than stand-alone genes that may not have any evidence of interactions thus increasing the chance of network effects and identifying therapeutic candidates AD. In STRING, each k-means cluster were characterized using biological processes identified by Gene Ontology ^36^. We then selected the central node(s) in each k-means cluster based on their connections with other genes in the network, with identified central node(s) having the highest number of connections (Figure 1). In other words, those with the most connections are preferentially chosen. Selecting highly interconnected gene nodes increases the likelihood of identifying candidates that are fundamentally important biologically relevant targets in AD.

After identifying the central nodes in each k-means cluster for the frontal cortex and cerebellum, respectively, we then undertook the second stage of our two-stage machine learning approach: supervised machine learning using decision trees (classification and regression trees (CART)) ^37^ (Figure 1). Here, the CART identifies which genes are best able to separate AD patients from healthy controls within the machine learning model. The use of CART has been well-established in large clinical and public health projects characterized by high-dimensional, heterogeneous data ^37, 38^. In the current study, the central gene nodes for each k-means cluster were used as features in the CART. The datasets for frontal cortex and cerebellum were each split into training (75%) and testing (25%) datasets. Training, tuning, and validating the model was done on the training dataset. Here, a 5-fold cross-validation was repeated three times to improve the accuracy estimates of the models ^39^. Cross-validation was used also as a tool for identifying the top performing gene predictors. The final evaluation of the CART model performance was done on the previously withheld testing dataset. CART was performed in R Studio v1.2.5033 (R v3.6.3) with libraries rpart, caret, and pROC.

### 2.4. Comparison of central node genes between frontal cortex and cerebellum

In parallel with our supervised machine learning analysis, we also sought to identify if there were any common central node genes between the frontal cortex and cerebellum. Identifying overlapping functional nodes is important to establish any common mechanisms underlying AD pathology in both regions. After identifying the overlap, we then performed a functional network analysis of these genes in STRING v11 to determine enriched pathways and biological processes that are common. We were also able to use the STRING analysis to confirm if any of the overlapping central node genes were themselves central nodes in the overlap functional network (Figure 1).

## 3. Results

### 3.1. Meta-analysis and unsupervised machine learning identifies novel dysregulated pathways and genes in AD

Unsupervised machine learning using principal component analysis (PCA) of the frontal cortex and cerebellum datasets demonstrated a clear clustering between the AD patients and healthy control groups along principal component (PC) 1 for the frontal cortex and PC2 for the cerebellum (Figure 2a and b).

**Figure 2.**
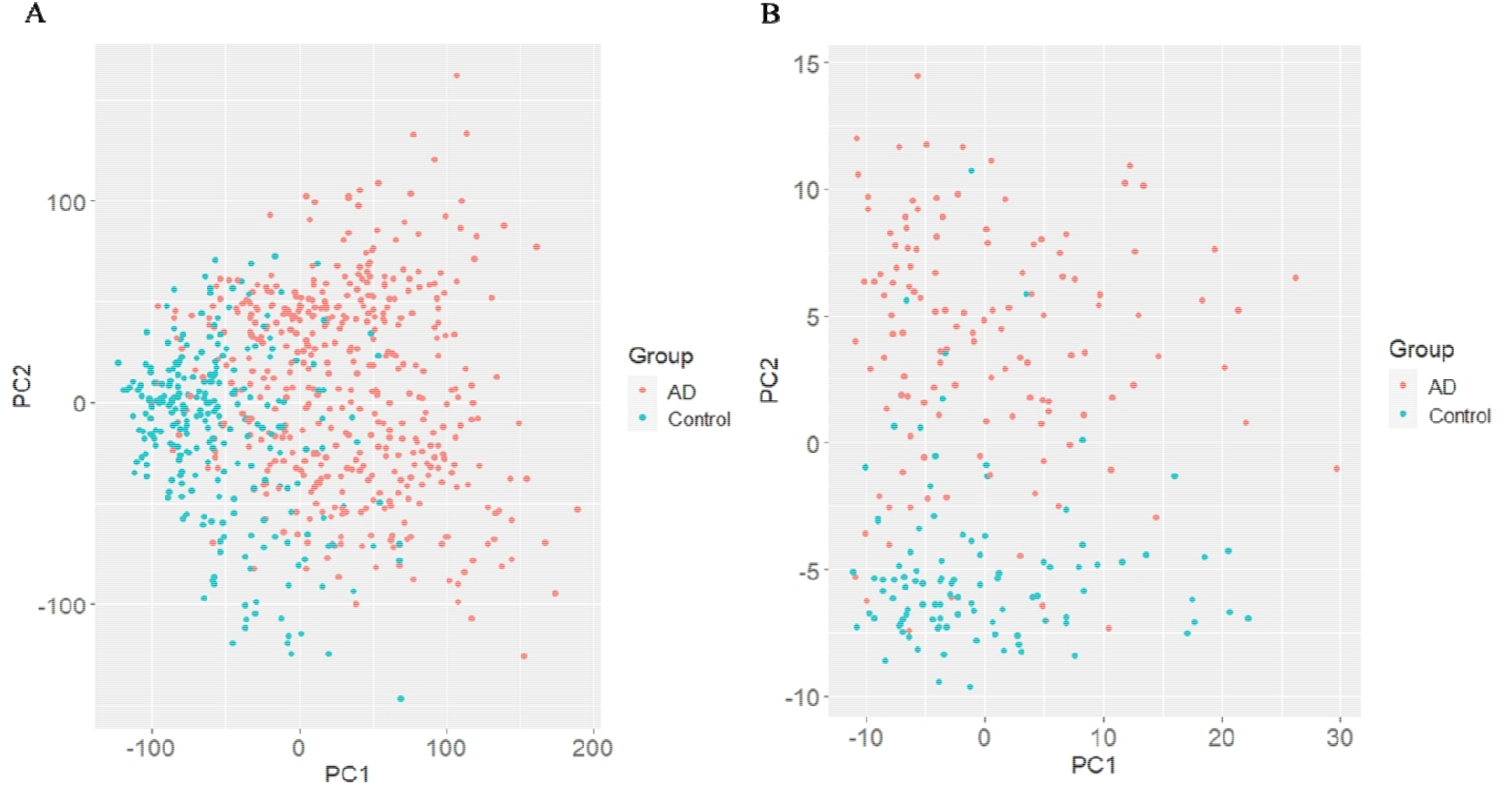
Principal component analysis of genes identified in the microarray meta-analysis reveals a clear between-group separation between the AD patients (red) and healthy controls (blue). **(a)** Frontal cortex. **(b)** Cerebellum.

Based on this finding, the top 1000 genes from frontal cortex PC1 and the top 1000 genes from cerebellum PC2 were identified as the most dysregulated genes in AD as they contributed the most to between-group variance (Supplementary Table 1). To determine the number of common pathways represented in these genes, a second unsupervised machine learning approach was performed using k-means clustering in STRING to identify genes in the network that have similar interconnections and overlapping pathways ^35^. Three k-means clusters were identified in both the frontal cortex and cerebellum (Figure 3). We then used Gene Ontology (GO) to characterize the biological functions of each of the clusters.

**Figure 3.**
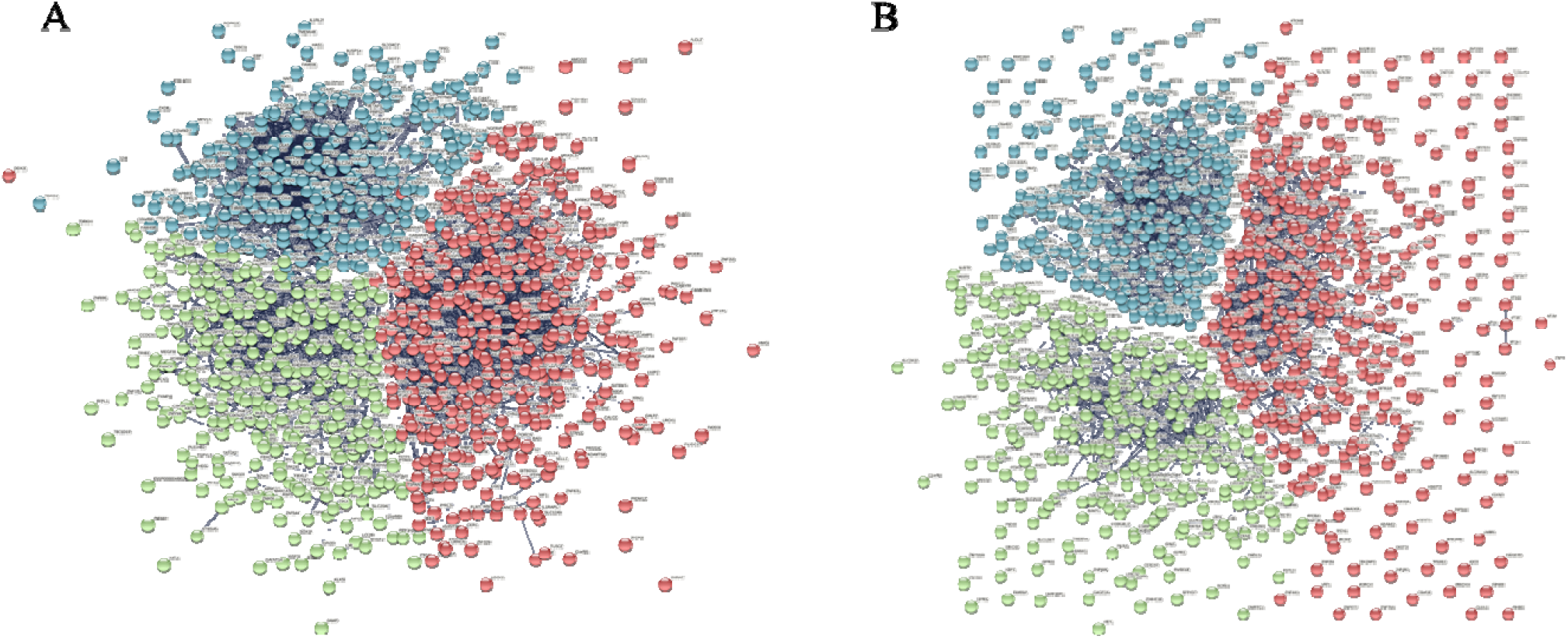
STRING k-means clustering of the top 1000 dysregulated genes in AD identified by the principal component analysis (PCA). **(a)** Frontal cortex principal component 1 (PC1) k-means clustering showed three distinct clusters of genes. Red n = 420, green n = 294, blue n = 286. **(b)** Cerebellum PC2 k-means clustering showed three distinct clusters of genes. Red n = 392, green n = 325, blue n = 283.

The first k-means cluster (red) for the frontal cortex was characterized by signaling processes (Supplementary Figure 1). Within this cluster, 15 central node genes were identified that play important roles in voltage-dependent calcium channels, guanine nucleotide-binding protein (G protein) formation and activity, AMPA receptor, cAMP-dependent protein kinase A (PKA), and the SNARE complex (Table 1; Supplementary Table 2). The second frontal cortical cluster (blue) was characterized by metabolic processes relating to macromolecules, proteins, and DNA (Supplementary Figure 2). Here, 26 genes were identified as being central nodes in the network with extensive roles in general cellular metabolic processes including DNA repair, transcriptional regulation, ubiquitin pathway regulation, and apoptosis (Table 1; Supplementary Table 2). The third k-means cluster identified in the frontal cortex (green) was related to mitochondrial processes. Within this cluster, there were two distinct sub-clusters identified based on biological function. The first was related to mitochondrial-specific energy, ATP, and oxidative phosphorylation and had 36 central nodes in the network (Supplementary Figure 3). These genes were all related to different mitochondrial subunits including: the F_0_-F_1_ ATP synthase (also known as mitochondrial complex V, ATP synthase), V-ATPase, mitochondrial complex I, and mitochondrial complex III (Table 1; Supplementary Table 2). The second sub-cluster within the green k-means cluster was defined by mitochondria-mediated cellular biosynthesis (Supplementary Figure 3) characterized by 10 central nodes where all genes were specific to the nuclear-encoded mitochondrial ribosomal 39S or 28S subunits, which play a central role in protein synthesis in mitochondria (Table 1; Supplementary Table 2).

**Table 1.** Summary of the overall characterization of the k-means clusters identified in the frontal cortex principal component 1 (PC1).

| **K-means Cluster** | **Functional Pathway Characterization** | **Number of Central Gene Nodes** | **Roles of Central Gene Nodes** |
| --- | --- | --- | --- |
| 1 (red) | Signaling processes | 15 | Voltage-dependent calcium channels, G-protein formation and activity, AMPA receptor, cAMP-dependent protein kinase A, and SNARE complex |
| 2 (blue) | Metabolic processes (macromolecular, proteins, and DNA) | 26 | General cellular metabolic processes, DNA repair, transcriptional regulation, ubiquitin pathway regulation, and apoptosis |
| 3A (green) | Mitochondria (energy, ATP, and oxidative phosphorylation) | 36 | ATP synthase, V-ATase, mitochondrial complex I, mitochondrial complex III |
| 3B (green) | Mitochondria (cellular biosynthesis) | 10 | Mitochondrial ribosomal 39S and 28S subunits, mitochondrial protein synthesis |

The characterization of the cerebellum k-means clusters revealed identical functional pathways to those in the frontal cortex (Table 2) albeit genes and the number of central gene nodes were different. The metabolic processes pathway was characterized by 17 central gene nodes which plays a role in transcription and nucleic acid binding (Table 2; Supplementary Figure 4; Supplementary Table 3). The signaling processes pathway had 10 central gene nodes (Supplementary Figure 5) and there was some overlap here in terms of the roles of the genes with the frontal cortex signaling processes pathway including voltage-dependent calcium channels and SNARE complexes. Unique to the cerebellum signaling pathway, however, was the involvement of cell-cell junctions, actin filament and cytoskeleton, and vesicular transport (Table 2; Supplementary Table 3). Like the frontal cortex, the third (green) k-means cluster identified in the cerebellum was related to mitochondrial processes and was subdivided into two distinct sub-clusters: energy, ATP, and oxidative phosphorylation and cellular biosynthesis (Supplementary Figure 6). The energy, ATP, and oxidative phosphorylation sub-cluster was characterized by 18 central gene nodes involved in ATP synthase, V-ATPase, and mitochondrial complexes I and III (Table 2; Supplementary Table 3). The cellular biosynthesis sub-cluster included 11 central gene nodes that made up components of the mitochondrial ribosomal 39S and 28S subunits and were involved in mitochondrial ribosomal protein synthesis (Table 2; Supplementary Table 3).

**Table 2.**
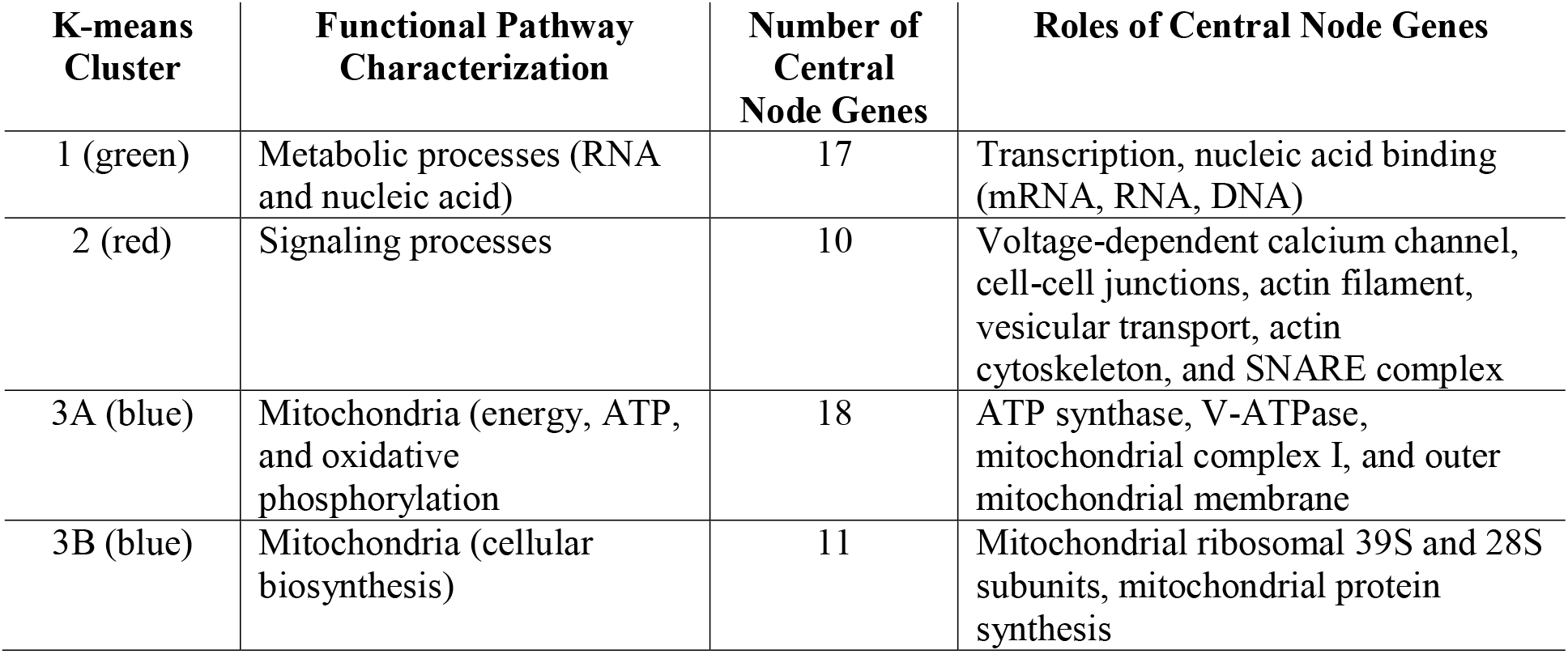
Summary of the overall characterization of the k-means clusters identified in the cerebellum across principal component (PC) 2.

| **K-means Cluster** | **Functional Pathway Characterization** | **Number of Central Node Genes** | **Roles of Central Node Genes** |
| --- | --- | --- | --- |
| 1 (green) | Metabolic processes (RNA and nucleic acid) | 17 | Transcription, nucleic acid binding (mRNA, RNA, DNA) |
| 2 (red) | Signaling processes | 10 | Voltage-dependent calcium channel, cell-cell junctions, actin filament, vesicular transport, actin cytoskeleton, and SNARE complex |
| 3A (blue) | Mitochondria (energy, ATP, and oxidative phosphorylation) | 18 | ATP synthase, V-ATPase, mitochondrial complex I, and outer mitochondrial membrane |
| 3B (blue) | Mitochondria (cellular biosynthesis) | 11 | Mitochondrial ribosomal 39S and 28S subunits, mitochondrial protein synthesis |

### 3.2. Supervised machine learning reveals the top signaling pathways for AD

The central gene nodes identified in the k-means clusters and sub-clusters within each brain structure were subsequently analyzed using supervised machine learning. A classification and regression tree (CART) algorithm was applied to each cluster, respectively, to compare the performance of the clusters in predicting AD. Performance indicators used included optimal sensitivity (correctly identifies AD patients), specificity (correctly identifies healthy controls), accuracy (correct number of classifications (AD / healthy control)) and AUC-ROC curve (capability of the model to distinguish between AD patients and healthy controls). The optimal performing CART was selected based on a high specificity and sensitivity as well as AUC.

Using these metrics, the top performing cluster for the frontal cortex was related to mitochondrial energy, ATP, and oxidative phosphorylation. This was followed very closely by the synaptic signaling, metabolic processes, and mitochondrial cellular biosynthesis clusters (Table 3, Figure 4). While sensitivity was the same between both models, the mitochondrial energy, ATP, and oxidative phosphorylation cluster model had a slightly higher specificity and AUC, suggesting that this model was slightly better at identifying controls (Figure 4).

**Table 3.** CART performance metrics for the k-means clusters and sub-clusters in the AD frontal cortex. AUC: area under the curve.

| Cluster | Specificity | Sensitivity | Accuracy | AUC |
| --- | --- | --- | --- | --- |
| Mitochondrial energy, ATP, and oxidative phosphorylation | 0.83 | 0.93 | 0.85 | 90.1 |
| Signaling | 0.83 | 0.92 | 0.86 | 86.6 |
| Metabolic processes | 0.82 | 0.91 | 0.84 | 88.2 |
| Mitochondrial cellular biosynthesis | 0.81 | 0.88 | 0.83 | 89.7 |

**Figure 4.**
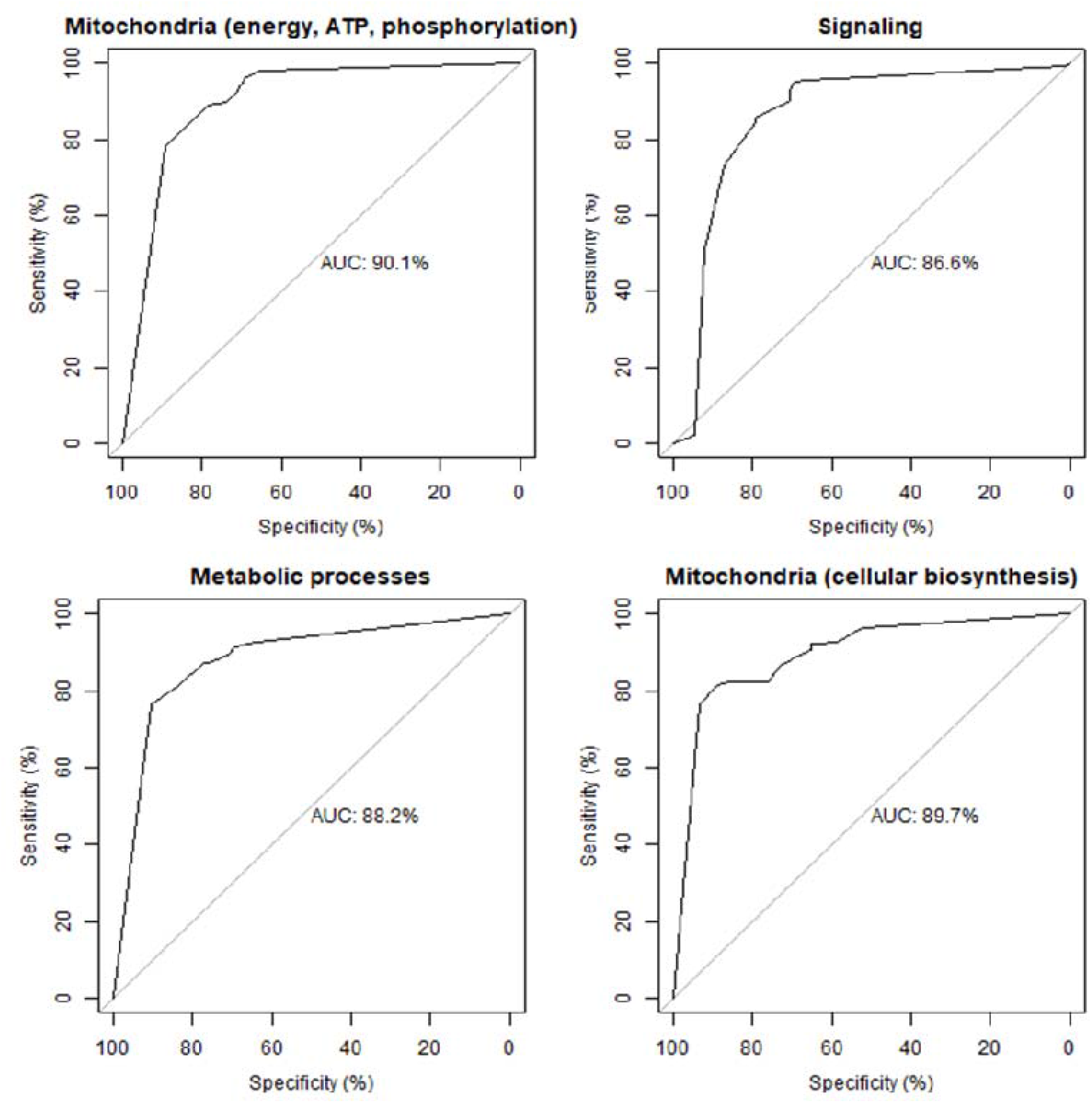
Receiver operating characteristic (ROC) curve of the CART models performance for each cluster in the frontal cortex. AUC: area under the curve.

For the cerebellum, the top performing cluster was for mitochondrial cellular biosynthesis. This was closely followed by metabolic processes and signaling and finally by mitochondrial energy, ATP, and oxidative phosphorylation (Table 4, Figure 5). Although the levels of sensitivity vary between pathways in each brain region, all four pathways were common to both brain regions.

**Table 4.** CART performance metrics for the k-means clusters and sub-clusters in the AD cerebellum.

| Cluster | Specificity | Sensitivity | Accuracy | AUC |
| --- | --- | --- | --- | --- |
| Mitochondrial cellular biosynthesis | 0.80 | 0.85 | 0.76 | 88.0 |
| Metabolic processes | 0.76 | 0.89 | 0.82 | 83.9 |
| Signaling | 0.74 | 0.89 | 0.81 | 81.0 |
| Mitochondrial energy, ATP, and oxidative phosphorylation | 0.67 | 0.86 | 0.76 | 85.4 |

**Figure 5.**
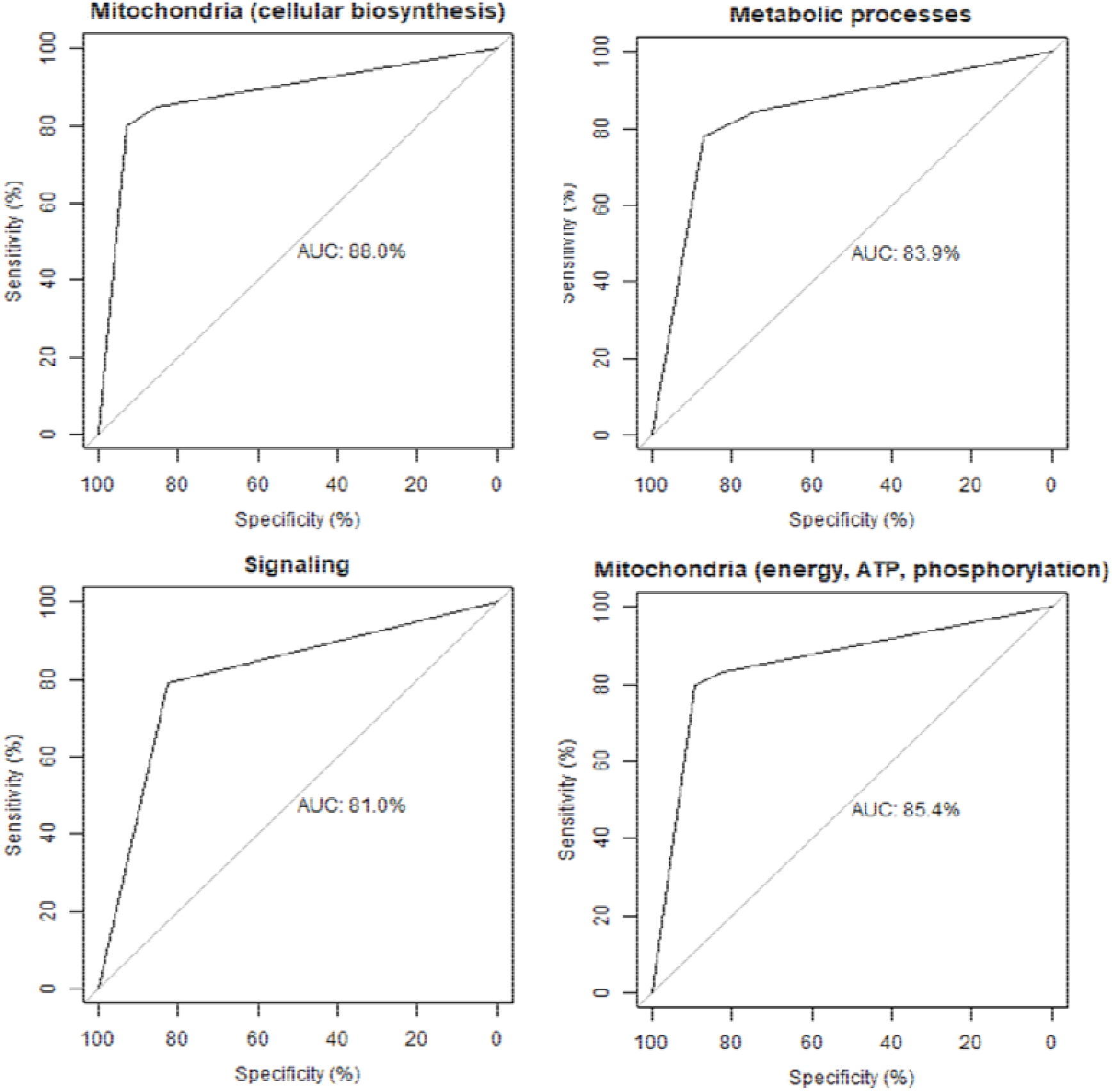
Receiver operating characteristic (ROC) curve of the CART models performance for each cluster in the cerebellum. AUC: area under the curve.

### 3.3. Analysis of the overlapping gene nodes between frontal cortex and cerebellum reveals the importance of ATP5L and ATP5H

We then identified the common central node genes in the four overlapping pathways, (Table 5). Overlapping central gene nodes were disproportionately represented from the mitochondrial energy, ATP, and oxidative phosphorylation pathway, with 10 genes being dysregulated in AD across the frontal cortex and cerebellum. These genes largely comprised subunits of ATP synthase (n = 5), followed by the V-ATPase (n = 2), mitochondrial complex I (n = 2), and mitochondrial complex III (n = 1). Three central gene nodes from mitochondrial cellular biosynthesis pathway: one 39S subunit ribosomal protein gene and two 28S subunit ribosomal protein genes. Only two central gene nodes overlapped in the signaling pathway between the cerebellum and frontal cortex: a subunit for a voltage-dependent calcium channel and a SNARE complex member (Table 5).

**Table 5.**
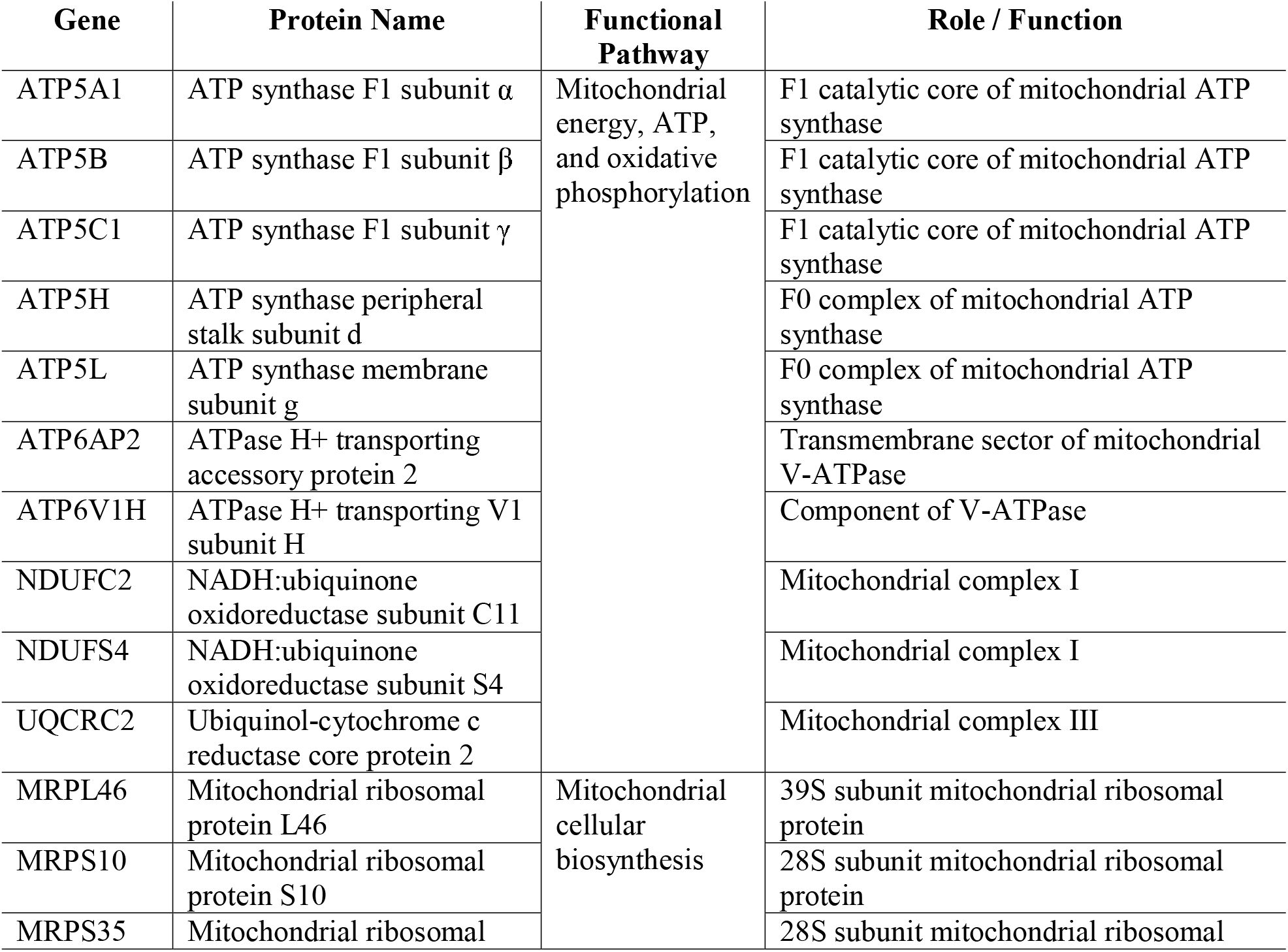
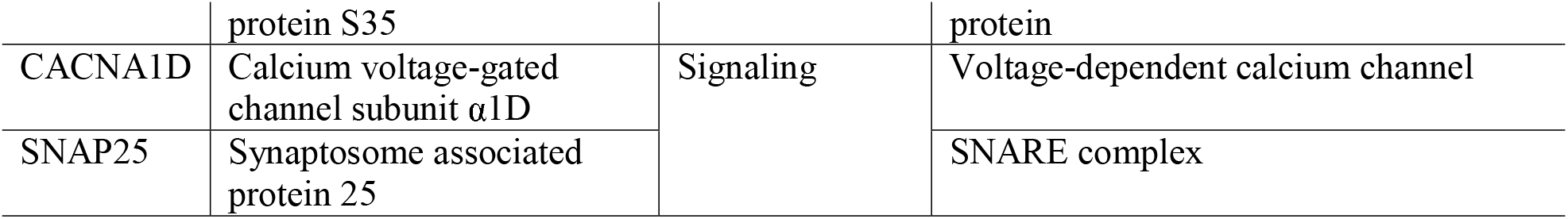
Overlapping central gene nodes between the frontal cortex and cerebellum.

Given these central gene nodes overlapped in both the cerebellum and frontal cortex, we then sought to identify if any of these genes were themselves central nodes (i.e. had the most connections) within a network using STRING. We found that two of these genes had the most connections with the others and therefore were likely central nodes within this sub-group. They were ATP5L and ATP5H, each with nine connections across the other mitochondrial energy, ATP, and oxidative phosphorylation genes (Figure 6). The biological processes that were enriched in this network included oxidative phosphorylation (FDR = 1.87e-10), mitochondrial ATP synthesis coupled proton transport (FDR = 3.50e-8), cristae formation (FDR = 9.98e-8), and mitochondrial transport (FDR = 6.76e-6).

**Figure 6.**
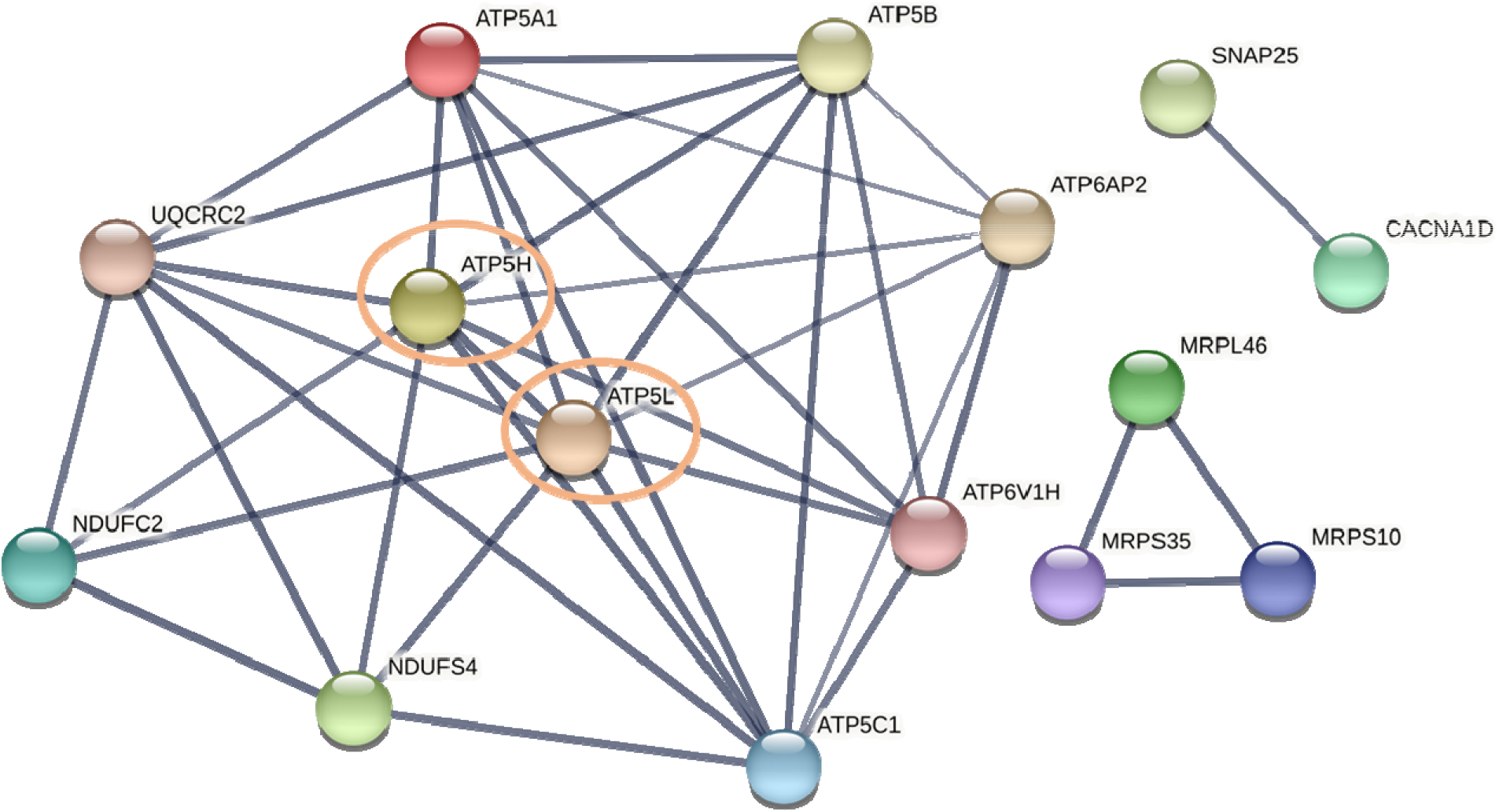
STRING functional network analysis of the overlapping central gene nodes between the frontal cortex and cerebellum of AD patients. The STRING network analysis includes the following interaction sources: experiments, databases, co-expression, neighborhood, and gene fusion. The minimum interaction score was set to 0.7 (high confidence). Thickness of the line indicates the confidence in the interaction.

## 4. Discussion

Identifying novel gene candidates in AD is challenging. We approached this challenge by performing a meta-analysis of microarray data from the frontal cortex and cerebellum of patients with AD. We then used an artificial intelligence-driven method, specifically a combination of unsupervised and supervised machine learning, to identify novel gene candidates for future study.

Across both the frontal cortex and cerebellum there were four functional pathways found to be dysfunctional in AD. These included 1) signaling, 2) metabolic processes, 3) mitochondrial energy, ATP, and oxidative phosphorylation, and 4) mitochondrial cellular biosynthesis. Despite the overlapping functional pathways between the two regions, there were pronounced differences in the relative importance of these pathways and the central gene nodes identified within each (Figure 6). First, the supervised machine learning analysis using CART identified that in the frontal cortex the mitochondrial energy, ATP, and oxidative phosphorylation pathway was the best predictor of AD. In the cerebellum, however, the mitochondrial cellular biosynthesis pathway was the best predictor of AD. There was little overlap (15/143) in the central gene nodes between the frontal cortex and cerebellum where fifteen central gene nodes were identical and most (10/15) were members of the mitochondrial energy, ATP, and oxidative phosphorylation functional pathway (Figure 5). A STRING functional network analysis of these overlapping genes found they were enriched in mitochondrial-related biological processes, including oxidative phosphorylation, mitochondrial ATP synthesis coupled proton transport, cristae formation, and mitochondrial transport. Further, two genes in this network were the most highly connected: ATP5L and ATP5H and therefore represented central gene nodes within this overlapping sub-network.

**Figure 5.**
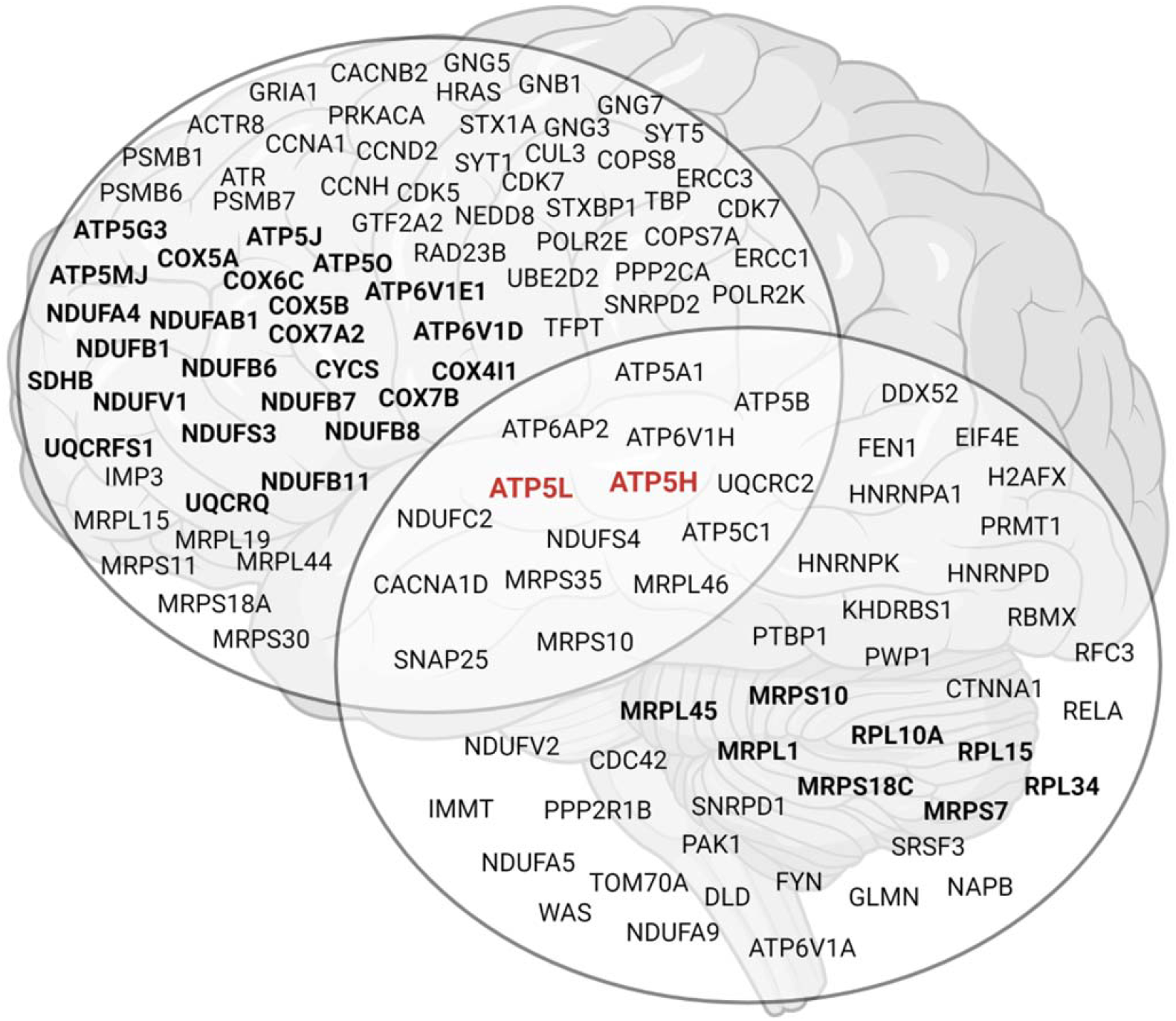
Summary of the central gene nodes identified in the frontal cortex and cerebellum, including the overlap in central gene nodes between the two regions. Black bolded genes represent those identified by the classification and regression trees (CART) as being significant predictors of AD. Red bolded genes represent the two central gene nodes of the overlapping genes between the frontal cortex and cerebellum.

Mitochondria are increasingly being shown to contribute to the development and progression of AD, with evidence for both primary and secondary dysfunctional mitochondrial cascades (for reviews see ^40, 41^). Specifically, mitochondrial dysfunction not only affects AD pathology, including APP activity and β amyloid (Aβ) accumulation, but AD pathology also leads to further mitochondrial dysfunction ^41^. Our findings are consistent with these studies showing that oxidative phosphorylation and mitochondrial ribosomal subunit mRNA are decreased in the bloods of patients with MCI who were at risk of developing AD as well as the AD patients themselves ^42, 43^. This highlights an early pattern of systemic mitochondrial dysfunction that both precedes AD pathology and persists across the disease trajectory. These findings are further confirmed by a recent proteome analysis of the AD brain tissue showing that dysregulated mitochondrial complexes, including ATP synthase, are potential drivers for AD pathology ^44^. The results of the present study also suggest that the role of the mitochondria in AD may be more nuanced. Specifically, our supervised machine learning analysis found that dysregulated oxidative phosphorylation and ATP synthesis genes are altered in the frontal cortex whereas dysregulated mitochondrial ribosomal genes were altered in the cerebellum. This suggests that although the mitochondria, broadly, are important, there may be key differences in specific mitochondrial mechanisms underlying pathology across brain regions. The reason underlying these differences, however, is unclear. It is also important to consider that this finding may be due to mitochondrial dysfunction resulting from varying levels of disease load across brain regions. For example, amyloid plaque deposition spreads to the cerebellum only in the end stages of AD ^45^ and perhaps amyloid β leads to mitochondrial ribosomal dysfunction. As discussed above, however, there is growing evidence that mitochondrial dysfunction precedes AD pathology. Future research would strongly benefit from examining these possibilities further.

Despite the finding of different mitochondrial mechanisms in the frontal cortex and cerebellum, we did find that two ATP synthase genes (ATP5L and ATP5H) represented a potential common mechanism underlying AD pathology in both regions. ATP5L encodes the g subunit and ATP5H encodes subunit d of the F_0_ membrane-spanning component of ATP synthase. ATP synthase, also known as mitochondrial complex V, is the final step in the oxidative phosphorylation pathway and the site of adenosine diphosphate (ADP) to adenosine triphosphate (ATP) conversion. It also plays a significant role in the formation of the mitochondrial inner membrane cristae ^46^. There has been some research into the role of ATP synthase in AD. Dysfunctional ATP synthase has been shown to sensitize mitochondrial permeability transition pore formation and lead to AD pathology, including amyloid β ^47^. AD pathology has also been found to lead to further decreases in ATP synthase activity via modifications to the α subunit of the F1 catalytic core ^46, 48, 49^. There is also some evidence that patients with AD have serum anti-ATP synthase β subunit autoantibodies, suggesting that mitochondrial dysfunction may be driven by autoimmunity ^50^. Despite the growing evidence that ATP synthase plays a role in AD, there have been no studies that specifically examine a role for the g (ATP5L) and d (ATP5H) subunits. Interestingly, however, two genome-wide association study (GWAS) meta-analysis of approximately 25,000 and 50,000 people, respectively, identified a shared ATP5H/KCTD2 locus for AD risk ^51, 52^. In line with this finding, a recent analysis of polygenic risk scores for patients with AD found that oxidative phosphorylation genes, including ATP synthase, were strongly associated with risk of AD ^53^. Given these genetic findings and our own that ATP5L and ATP5H dysregulation represents a possible brain-wide mechanism in AD pathology, future research would benefit from studying these ATP synthase subunits further.

Although our work presents a strong case for a key role of novel genes in AD, there are some limitations that should be taken into consideration. First, based on our inclusion and exclusion criteria, we only identified three datasets that were appropriate for inclusion in the current study: two from the frontal cortex and one from the cerebellum. Although we did seek to include data from other brain regions, including the hippocampus and entorhinal cortex, the microarray datasets for these did not have the sample size to support our machine learning-based analyses. As such, we are unable to include a comparison of the genes and pathways that are involved in these regions to those discussed here. Future research would benefit from focusing on expanding gene expression data for these regions to enable large-scale analyses and the application of artificial intelligence methods. Our current findings, however, present a strong case for key overlapping genes across the frontal cortex and cerebellum despite differences in AD pathophysiology ^54^, which highlight the possibility of common underlying mechanisms. A second consideration is that our study has used established databases (e.g. STRING ^35^, Gene Ontology ^36^) to identify functional networks and pathways in AD. An inherent limitation of these databases is that they rely on existing interactions between proteins and networks that have been previously identified in experimental literature. As such, the use of these databases precludes the possibility of other central gene node(s) that do not yet have a wealth of experimental data. Although there is no way to overcome this limitation, we strongly suggest that future experimental work confirm the findings presented here.

In summary, our study reports an artificial intelligence-driven identification of novel gene candidates in the frontal cortex and cerebellum of patients with AD using a relatively unbiased methodology. Our findings highlight the importance of nuclear-encoded mitochondrial genes involved in oxidative phosphorylation in the frontal cortex and mitochondrial ribosomal protein synthesis in the cerebellum. Further, we found ATP synthase function as a possible common mechanism underlying AD pathology across brain regions. Together, these findings highlight the possibility that mitochondrial dysfunction and pathophysiological mechanisms in AD may be brain region specific and that ATP synthase subunit dysregulation is a common mechanism. These candidates should be investigated further as they may have significant implications for understanding the etiology of AD and future therapeutic strategies.

## Data Availability

All data produced in the present study are available upon reasonable request to the authors

## Data Availability Statement

This meta-analysis did not generate any new data and used three publicly accessible datasets from the Gene Expression Omnibus database (GSE44770, GSE33000, and GSE44768). The top 1000 genes identified by the principal component analyses are available in Supplementary Table 1.

## Contributions

C.A.F. and A.S. conceived the study, acquired funding, performed the machine learning and STRING network analyses, and made the figures. C.A.F. curated the data for analysis. C.A.F., F.D., W.A.G., D.A.B., and A.S. contributed to the methodology and wrote, read, and approved the final manuscript.

## Acknowledgements

This work was supported by a Macquarie University Research Acceleration Scheme grant to C.A.F. and A.S. (# 5224420).

