## Supplementary Figures for "Artificial intelligence-driven meta-analysis of brain gene expression identifies novel gene candidates and a role for mitochondria in Alzheimer’s Disease"

*Authors to whom correspondence should be addressed.

**Supplementary Figures**

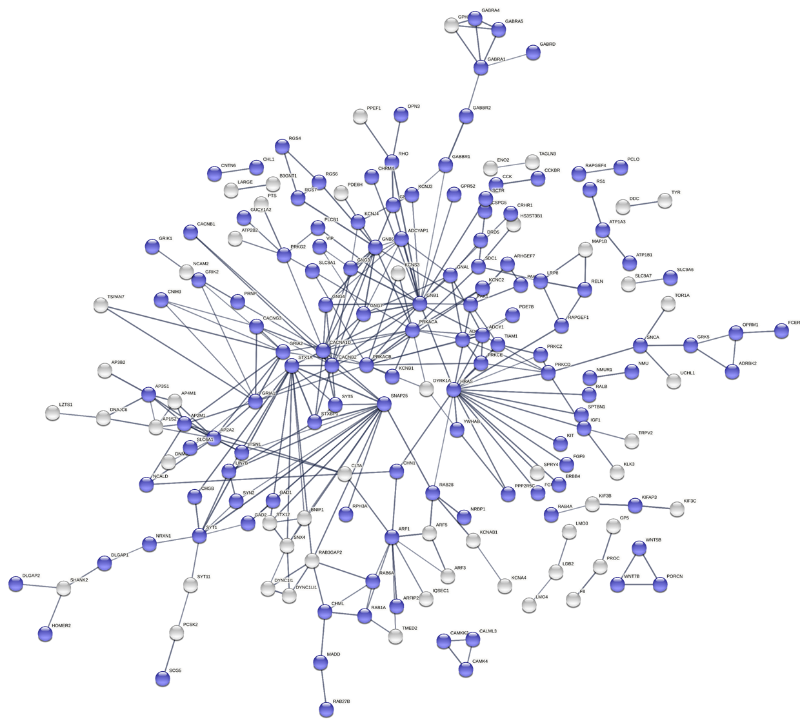

**Supplementary Figure 1**. Characterization of the biological processes of frontal cortex principal component 1 (PC1) k-means cluster 1 using Gene Ontology and STRING. The STRING network analysis includes the following interaction sources: experiments, databases, co-expression, neighborhood, and gene fusion. The minimum interaction score was set to 0.7 (high confidence) and disconnected nodes in the network are hidden. Thickness of the line indicates the confidence in the interaction. The blue nodes indicate genes involved in signaling (FDR = 1.68e-26).

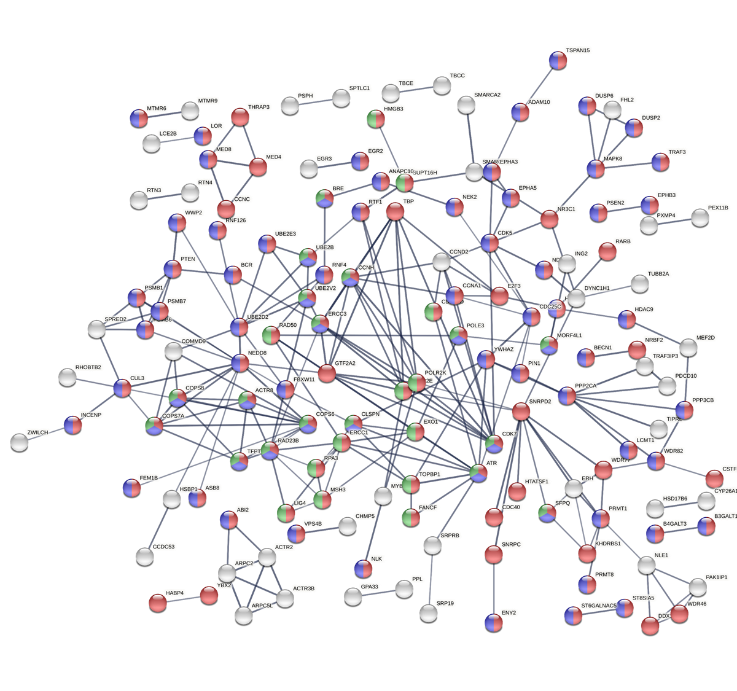

**Supplementary Figure 2**. Characterization of the biological processes of frontal cortex principal component 1 (PC1) k-means cluster 2 using Gene Ontology and STRING. The STRING network analysis includes the following interaction sources: experiments, databases, co-expression, neighborhood, and gene fusion. The minimum interaction score was set to 0.7 (high confidence) and disconnected nodes in the network are hidden. Thickness of the line indicates the confidence in the interaction. Red nodes indicate genes involved in macromolecule metabolic processes (FDR = 2.09e-13). Blue nodes indicate genes involved in protein metabolic processes (FDR = 5.22e-9). Green nodes indicate genes involved in DNA metabolic processes (FDR = 5.14e-6).

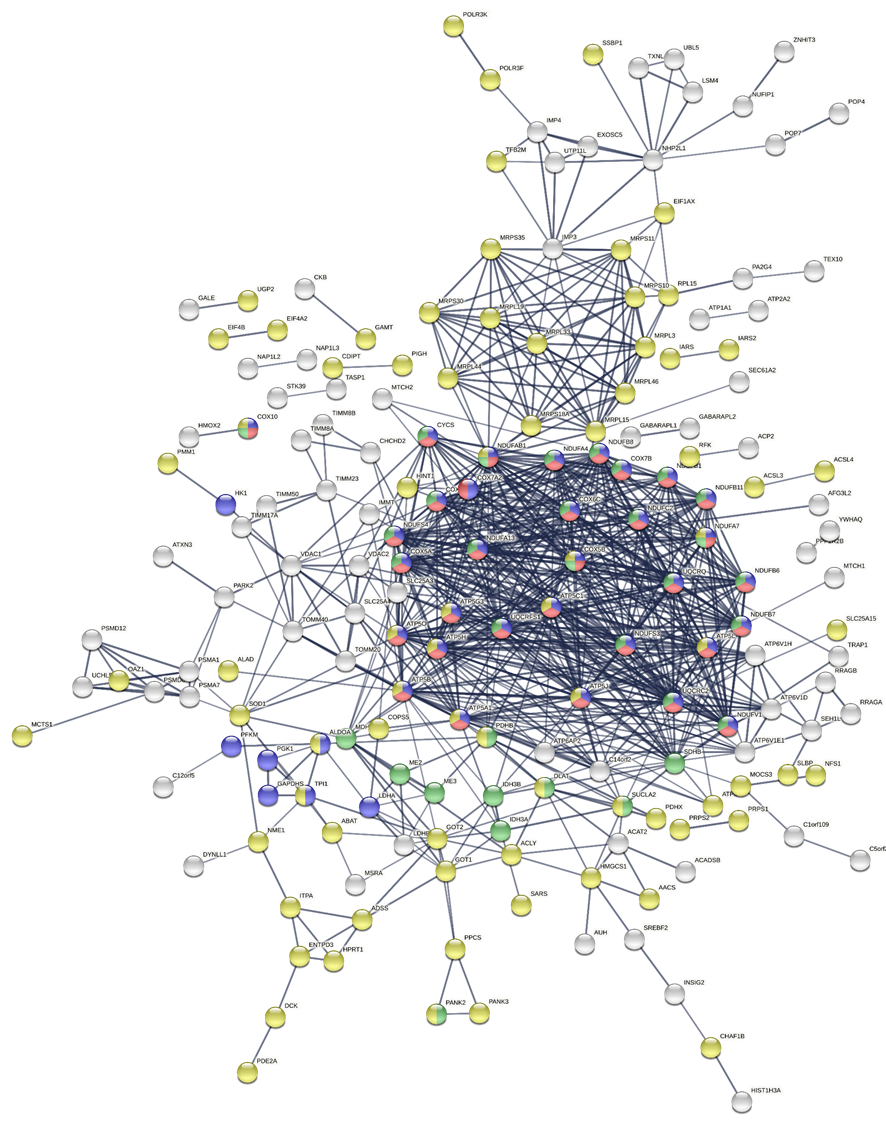

**Supplementary Figure 3**. Characterization of the biological processes of frontal cortex principal component 1 (PC1) k-means cluster 3 using Gene Ontology and STRING. The STRING network analysis includes the following interaction sources: experiments, databases, co-expression, neighborhood, and gene fusion. The minimum interaction score was set to 0.7 (high confidence) and disconnected nodes in the network are hidden. Thickness of the line indicates the confidence in the interaction. Blue nodes indicate genes involved in ATP metabolic processes (FDR = 4.75e-26). Red nodes indicate genes involved in oxidative phosphorylation (FDR = 2.76e-24). Green nodes indicate genes involved in cellular respiration (FDR = 2.76e-24). Together, the blue, red, and green nodes form sub-cluster 3A: mitochondria (ATP, energy, and oxidative phosphorylation). Yellow nodes indicate genes involved in cellular biosynthetic processes (FDR = 2.01e-12) and form sub-cluster 3B: mitochondria (cellular biosynthesis).

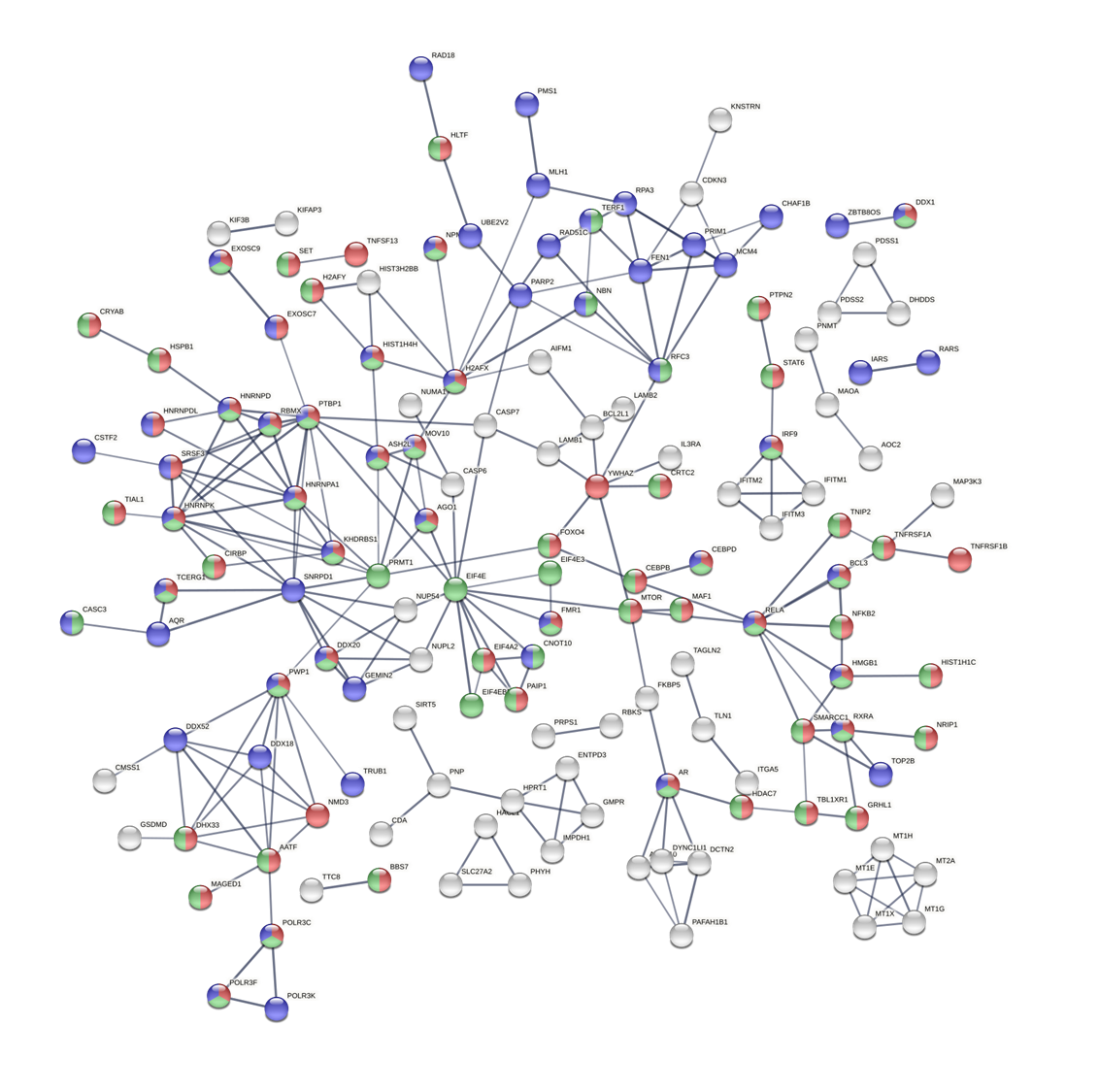

**Supplementary Figure 4**. Characterization of the biological processes of cerebellum principal component 2 (PC2) k-means cluster 1 using Gene Ontology and STRING. The STRING network analysis includes the following interaction sources: experiments, databases, co-expression, neighborhood, and gene fusion. The minimum interaction score was set to 0.7 (high confidence) and disconnected nodes in the network are hidden. Thickness of the line indicates the confidence in the interaction. Green nodes indicate genes involved in regulation of cellular biosynthetic processes (FDR = 5.54e-7). Red nodes indicate genes involved in regulation of RNA metabolic processes (FDR = 9.90e-7). Blue nodes indicate genes involved in nucleic acid metabolic processes (FDR = 0.00037).

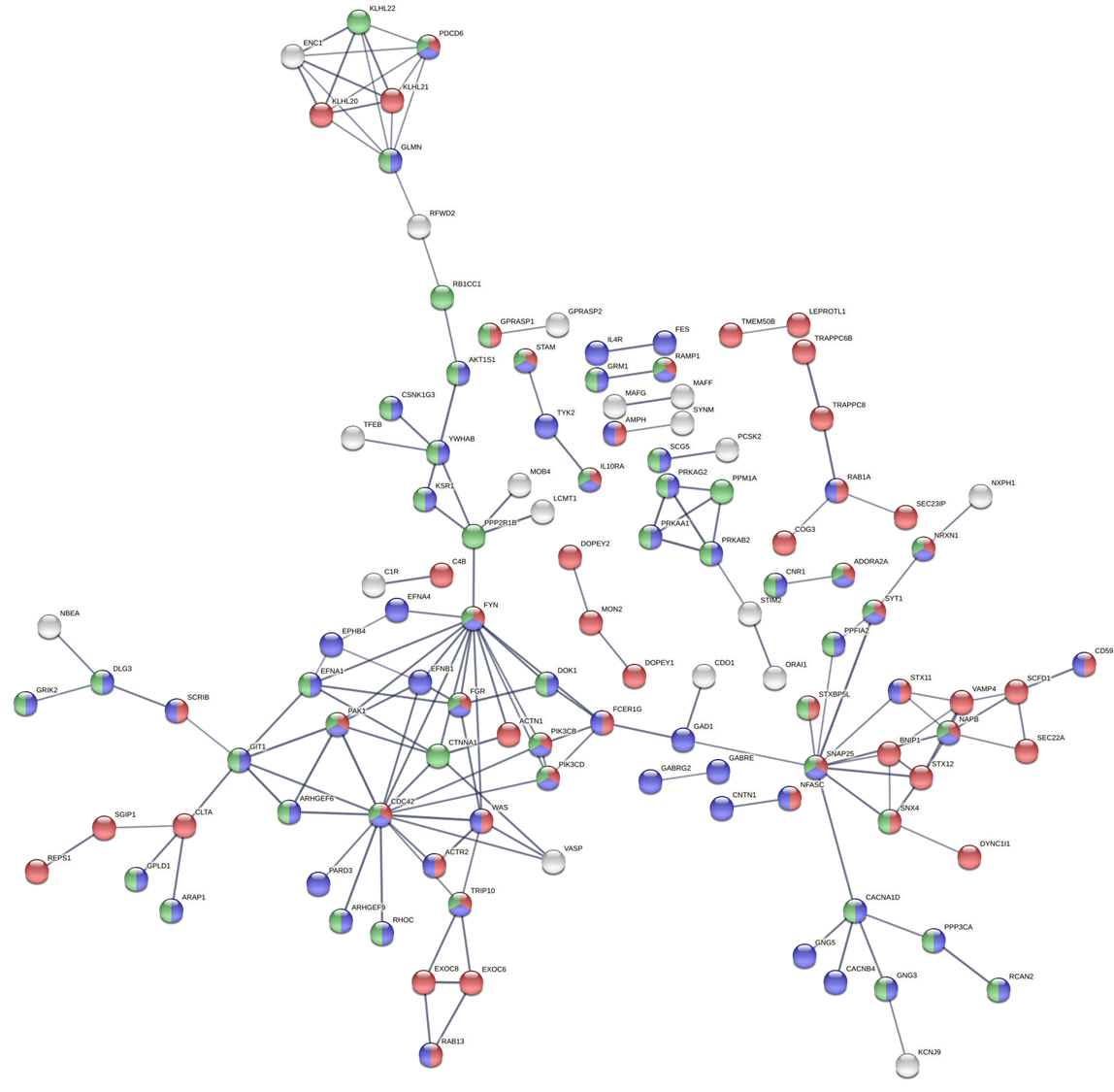

**Supplementary Figure 5**. Characterization of the biological processes of cerebellum principal component 2 (PC2) k-means cluster 2 using Gene Ontology and STRING. The STRING network analysis includes the following interaction sources: experiments, databases, co-expression, neighborhood, and gene fusion. The minimum interaction score was set to 0.7 (high confidence) and disconnected nodes in the network are hidden. Thickness of the line indicates the confidence in the interaction. Red nodes indicate genes involved in vesicle-mediated transport (FDR = 7.81e-13). Blue nodes indicate genes involved in signaling (FDR = 2.42e-5). Green nodes indicate genes involved in regulation of signaling (FDR = 6.62e-5).

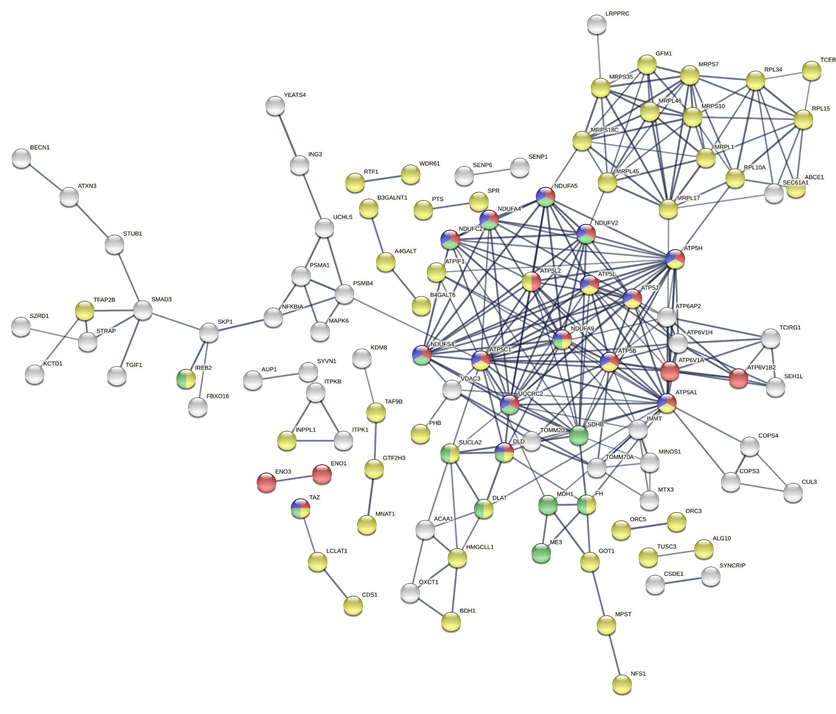

**Supplementary Figure 6**. Characterization of the biological processes of cerebellum principal component 2 (PC2) k-means cluster 3 using Gene Ontology and STRING. The STRING network analysis includes the following interaction sources: experiments, databases, co-expression, neighborhood, and gene fusion. The minimum interaction score was set to 0.7 (high confidence) and disconnected nodes in the network are hidden. Thickness of the line indicates the confidence in the interaction. Red nodes indicate genes involved in ATP metabolic processes (FDR = 7.04e-9). Blue nodes indicate genes involved in oxidative phosphorylation (FDR = 3.31e-7). Green nodes indicate genes involved in cellular respiration (FDR = 2.69e-7). Together, the blue, red, and green nodes form subcluster 3A: mitochondria (ATP, energy, and oxidative phosphorylation). Yellow nodes indicate genes involved in cellular biosynthetic processes (FDR = 1.02e-8) and form sub-cluster 3B: mitochondria (cellular biosynthesis).
